# A school-based trauma-informed care and social-emotional learning intervention for adolescents with adverse childhood experiences in India: Protocol for a sequential mixed-methods study with a single-arm quasi-experimental evaluation

**DOI:** 10.64898/2026.10.02.26364571

**Authors:** Bhavneet Bharti, Nidhi Malhotra, Amrit Virk, Deepika Lamba, Chandni Pandey, Prahbhjot Malhi, Sahul Bharti

**Author notes:** Corresponding author Dr Bhavneet Bharti, Professor of Pediatrics and Head Social Pediatrics, Department of Pediatrics, Postgraduate Institute of Medical Education and Research (PGIMER), Sector 12, Chandigarh 160012, India.

## Abstract

**Background:** Adverse childhood experiences are major determinants of mental disorders, health-risk behaviours and non-communicable diseases across the life course, but evidence from India remains limited. This study aims to estimate the prevalence and patterns of adverse childhood experiences among Indian adolescents, examine their associations with mental health and non-communicable disease risk factors, and evaluate a culturally adapted school-based intervention integrating trauma-informed care and social-emotional learning.

**Methods:** A sequential mixed-methods study will be conducted among approximately 1,632 adolescents in 16 public and private schools in Chandigarh and Mohali, India. Phase I is a cross-sectional survey of adverse childhood experiences, mental health, perceived stress, sleep, screen time and behavioural and biological risk factors for non-communicable diseases. Phase II comprises focus group discussions and key informant interviews with students, parents, teachers, counsellors and mental health professionals to inform intervention development.

In Phase III, findings from both phases will guide development of the intervention using the ADDIE (Analysis, Design, Development, Implementation and Evaluation) instructional design framework.

In Phase IV, adolescents reporting ACE exposure will participate in a school-based intervention delivered over 8–12 weeks by trained psychologists, comprising trauma-informed social-emotional learning sessions, individual counselling sessions for high-risk adolescents (at least six sessions), school-wide awareness activities and parental sensitization. Effectiveness will be evaluated using a quasi-experimental pre–post design. The primary outcome will be change in emotional and behavioural difficulties measured by the Strengths and Difficulties Questionnaire (SDQ). Secondary outcomes will include changes in depressive symptoms (Patient Health Questionnaire-9), perceived stress, sleep quality, school functioning, and behavioural risk factors for NCDs. Recruitment began in April 2026 and is ongoing; data collection is expected to be complete by June 2028, and no results have yet been generated.

**Trial Registration:** Clinical Trials Registry–India, CTRI/2025/10/095615 (prospectively registered 6 October 2025).

Findings will be disseminated through peer-reviewed publications, policy briefs, and stakeholder workshops.

## Background

Adverse Childhood Experiences (ACEs) typically classified as abuse, neglect, and household dysfunction, are increasingly recognized as major determinants of health across the life course. Recent systematic reviews and meta-analyses underscore the widespread burden of adverse childhood experiences (ACEs), with 58% of children and adolescents globally experiencing at least one ACE during childhood. [1,2] In a comprehensive meta-analysis of 65 studies and 4,90,423 participants in 18 countries, Madigan et al. reported that 42% of participants had no history of ACE exposure, whereas 22%, 13%, and 8% experienced one, two, and three ACEs, respectively. Notably, approximately 15% of individuals had been exposed to four or more ACEs, a threshold consistently associated with elevated risks of adverse mental and physical health outcomes. [1] Evidence from several longitudinal studies including the landmark CDC-Kaiser ACE study [3] demonstrates that childhood adversity is associated with a wide range of adverse outcomes, including depression, anxiety, substance use, cancer, chronic pain, cardiometabolic diseases, poor educational attainment, homelessness and premature mortality. The cumulative burden of ACEs has been shown to exert a dose response effect with increasing exposure being associated with progressively higher life course risks of mental health problems and noncommunicable diseases (NCDs) in adulthood. [4]

However, studies evaluating Indian adolescents are limited highlighting a substantial evidence gap and the urgent need for context-specific research in a country that is home to the world’s largest adolescent population, particularly in the backdrop of growing burden of mental health problems and noncommunicable disease risk factors. Comprehensive data from school settings, particularly in India, remains scarce. This lack of evidence constrains understanding of the prevalence, patterns, and health consequences of ACEs in Indian adolescents and highlights the need for robust, locally generated research to inform policy and intervention development. On a national level, the proposed study aligns closely with the priorities outlined in the National Education Policy (NEP 2020) [5] and the National Mental Health Policy (2014) [6], both of which advocate for early, integrative, and preventive mental health strategies within educational settings. Current school-based health programs in India primarily focus on physical health and life skills with limited attention to trauma-informed care or ACE prevention.

This gap is particularly concerning given that schools provide a strategic platform for early identification, intervention and prevention strategies. By integrating trauma-informed care with Social-Emotional Learning (SEL), this research not only contributes to the body of scientific knowledge but also builds technical capacity among school professionals. Enhancing the ability of teachers and counsellors to recognize and respond to ACEs will help bridge the gap between policy intent and practical implementation, thereby strengthening school-based health programs. In India, where healthcare resources are already strained, preventing and mitigating ACE impacts through early intervention could yield significant public health and economic benefits.

In summary, the proposed study seeks to generate context-specific evidence on the prevalence and impact of ACEs among adolescents in North India and to develop a culturally appropriate trauma-informed Social-Emotional Learning intervention. By examining the relationships between ACE exposure, mental health outcomes, and behavioural risk factors for NCDs, the study will contribute to a deeper understanding of adolescent wellbeing and support the development of scalable, preventive strategies. The findings will have implications for educational institutions, health systems, and policymakers working to promote resilience, improve mental health, and reduce the long-term burden of trauma-related health inequities. The schedule of enrolment, interventions and assessments for the study is summarised in Fig 1.

**Fig 1.**
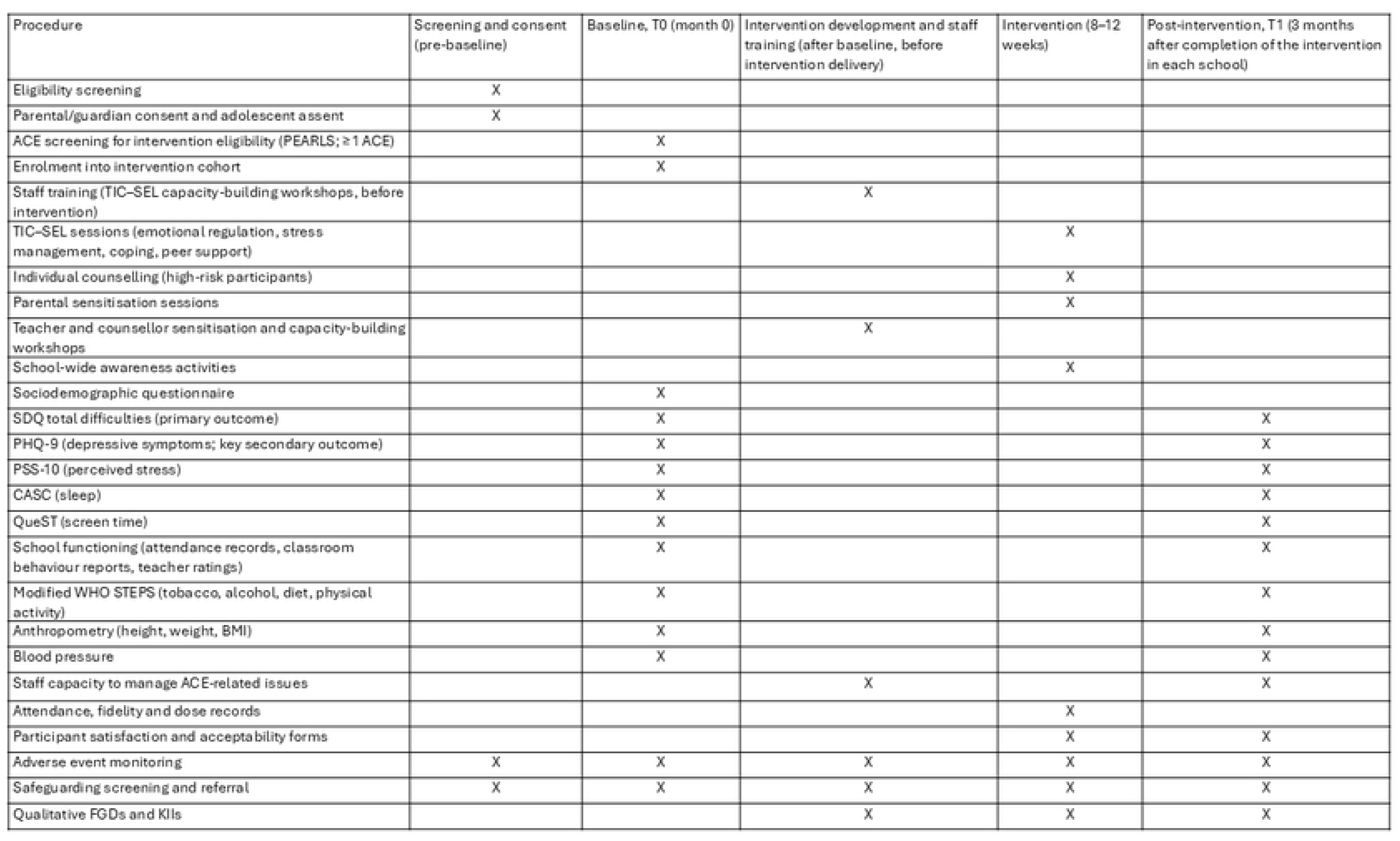
Schedule of enrolment, interventions and assessments (SPIRIT diagram). Time points are screening and consent (pre-baseline), baseline T0 (month 0), intervention development and staff training (after baseline and before intervention delivery), the intervention period (8–12 weeks) and post-intervention assessment T1 (3 months after completion of the intervention in each school). Staff capacity to manage ACE-related issues is self-rated by teachers and counsellors on a 0–10 Likert scale before and after training and at T1.

### Conceptual Framework and Theory of Change

The study is guided by the Adverse Childhood Experiences (ACEs) framework, the toxic stress model, and principles of Trauma-Informed Care (TIC). [7,8] The framework proposes that exposure to ACEs, including abuse, neglect, and household dysfunction, activates chronic stress-response pathways, resulting in toxic stress. Persistent activation of the hypothalamic-pituitary-adrenal (HPA) axis, autonomic nervous system dysregulation, and inflammatory processes can adversely affect neurodevelopment, emotional regulation, behaviour, and physical health. The conceptual framework and theory of change are summarised in Fig 2.

**Fig 2.**
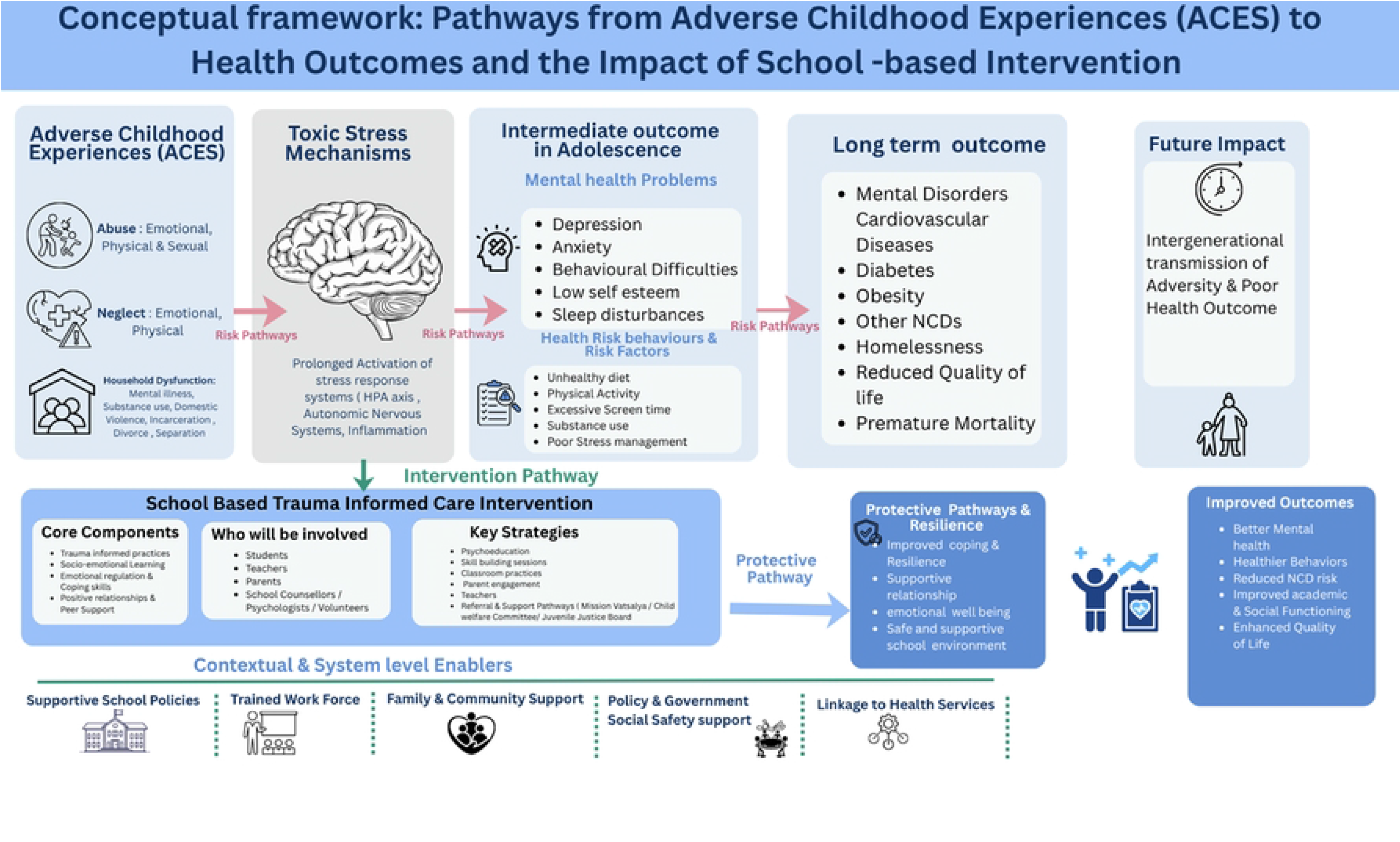
Conceptual framework illustrating the pathways linking adverse childhood experiences (ACEs) to toxic stress, neurodevelopmental and psychosocial consequences, health-risk behaviours, and long-term mental and physical health outcomes. The framework also depicts how a trauma-informed, school-based social-emotional learning intervention may strengthen resilience and protective factors, thereby mitigate the effects of adversity and improve adolescent wellbeing, academic functioning, and future health outcomes.

These effects may manifest during adolescence as depression, anxiety, behavioural difficulties, low self-esteem, sleep disturbances, and engagement in health-risk behaviours such as poor diet, physical inactivity, substance use, and maladaptive coping. Over time, these pathways increase the risk of mental disorders, noncommunicable diseases (NCDs), reduced quality of life, and intergenerational transmission of adversity.

The theory of change underpinning the intervention posits that early identification of ACEs, combined with a school-based trauma-informed Social-Emotional Learning (SEL) intervention, can interrupt these adverse pathways. [9] The intervention aims to enhance awareness of trauma, strengthen emotional regulation and coping skills, foster supportive peer and teacher relationships, engage parents, and establish referral pathways for students requiring additional support.

By creating safe, supportive, and trauma-sensitive school environments, the intervention is expected to strengthen resilience, self-efficacy, social connectedness, and emotional wellbeing. These protective factors are hypothesized to buffer the effects of toxic stress, reduce psychological distress and behavioural risk factors, and improve mental health, academic functioning, health behaviours, and overall quality of life. The effectiveness and sustainability of the intervention will be supported by enabling factors such as trained teachers, supportive school policies, family engagement, community participation, and linkages with health services.

Thus, the study hypothesizes that trauma-informed school interventions can transform adversity into resilience by modifying the pathways through which ACEs influence adolescent mental health and future NCD risk.

## Methodology

### Study Aims and Hypotheses

The primary aim of this study is to determine the prevalence and impact of Adverse Childhood Experiences (ACEs) among school-going adolescents and to develop, implement, and evaluate a culturally adapted school-based Trauma-Informed Care and Social-Emotional Learning (TIC–SEL) intervention to mitigate their adverse effects on mental health and behavioural risk factors for non-communicable diseases (NCDs).

We hypothesize that adolescents participating in the TIC–SEL intervention will demonstrate significant improvements in emotional and behavioural wellbeing following the intervention, as measured by reductions in Strengths and Difficulties Questionnaire (SDQ) scores.

### Objectives

**Objective 1:** To determine the prevalence and patterns of ACEs among school-going adolescents in Chandigarh and Mohali.

**Objective 2:** To determine the association between ACEs, mental health issues and bio-behavioural risk factors for NCDs among school-going adolescents, specifically:

2a. to examine the association between ACE exposure and mental health outcomes, including emotional and behavioural difficulties, depressive symptoms, perceived stress, sleep disturbances, and psychosocial wellbeing; and

2b. to examine the association between ACE exposure and behavioural and biological risk factors for NCDs, including unhealthy dietary practices, physical inactivity, screen-time behaviours, sleep problems, substance use, overweight/obesity, and elevated blood pressure.

**Objective 3:** To develop, implement and evaluate a targeted school-based Trauma-Informed Care and Social-Emotional Learning (TIC–SEL) intervention aimed at mitigating the impact of ACEs, specifically:

3a. to develop a culturally appropriate TIC–SEL intervention using quantitative findings, stakeholder perspectives, and the ADDIE instructional design framework;

3b. to evaluate the effectiveness of the intervention in improving emotional and behavioural wellbeing (SDQ; primary outcome) among adolescents exposed to ACEs;

3c. to assess the effectiveness of the intervention on secondary outcomes, including depressive symptoms (PHQ-9), perceived stress, sleep quality, school functioning, capacity of teachers and school counsellors to manage ACE-related issues, and behavioural risk factors for NCDs;

3d. to assess the acceptability, feasibility, fidelity, and implementation processes of the intervention among students, teachers, counsellors, and parents;

3e. to explore contextual facilitators and barriers influencing the implementation and sustainability of trauma-informed school interventions; and

3f. (exploratory) to explore differential effects of the intervention according to gender, age group, socioeconomic status, school type (public/private), and locality (urban/rural-peri-urban).

### Study Hypotheses

#### Research question

In school-going adolescents aged 11–15 years in Chandigarh and Mohali (Population), does a targeted school-based intervention integrating Trauma-Informed Care and Social-Emotional Learning (Intervention), compared with the adolescents’ own pre-intervention status (Comparison), reduce mental health difficulties and bio-behavioural risk factors for NCDs associated with ACEs, and strengthen the capacity of teachers and school counsellors to manage ACE-related issues (Outcome)? The following hypotheses were derived from this research question.

#### Primary Hypothesis

Adolescents exposed to ACEs who participate in the TIC–SEL intervention will demonstrate a significant reduction in emotional and behavioural difficulties, reflected by lower SDQ scores at post-intervention assessment compared with baseline (Objective 3b).

#### Secondary Hypotheses

1. Higher ACE exposure will be associated with poorer mental health outcomes, including increased depressive symptoms, perceived stress, emotional and behavioural difficulties, and sleep problems (Objective 2a).
2. Higher ACE exposure will be associated with a greater prevalence of behavioural and biological risk factors for NCDs (Objective 2b).
3. Adolescents receiving the Trauma Informed care–Socio Emotional Learning (TIC–SEL) intervention will demonstrate significant improvements in depressive symptoms, perceived stress, sleep quality, and school functioning (Objective 3c).
4. The TIC–SEL intervention will be acceptable, feasible, and implementable within school settings and will strengthen the capacity of teachers and school counsellors to identify and respond to ACE-related concerns (Objectives 3c and 3d).

### Study Design

This study will employ a sequential mixed-methods design conducted in four interrelated phases to comprehensively examine the prevalence and impact of Adverse Childhood Experiences (ACEs) among school-going adolescents and to develop and evaluate a culturally adapted school-based intervention. The overall study design is summarised in Fig 3.

**Fig 3.**
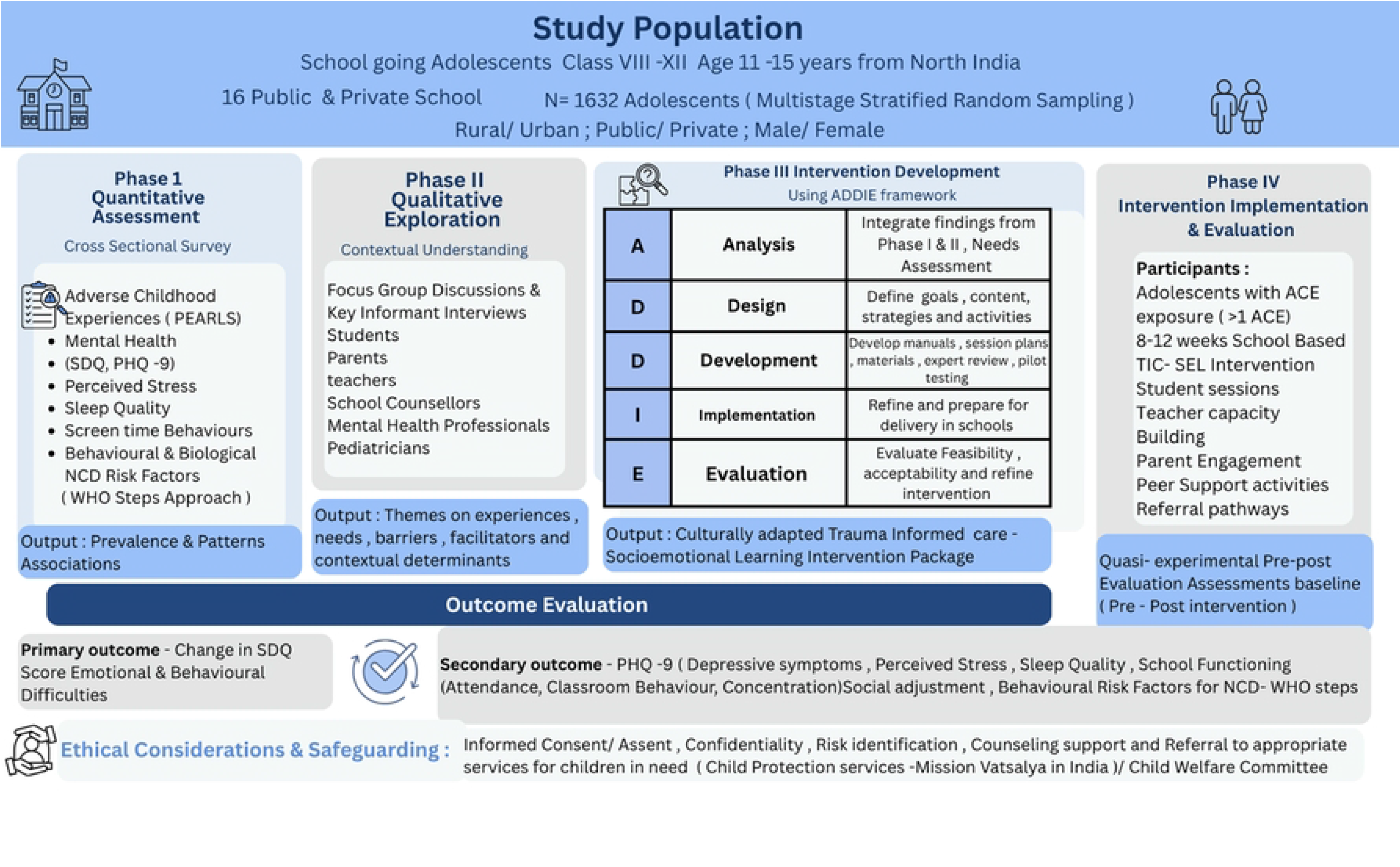
Sequential mixed-methods study design showing quantitative assessment, qualitative exploration, development of a culturally adapted TIC–SEL intervention using the ADDIE framework, and quasi-experimental evaluation among adolescents with ACE exposure. Primary outcome is change in SDQ scores; secondary outcomes include depression, perceived stress, sleep quality, school functioning, and NCD risk behaviours. Safeguarding procedures, counselling support, and referral pathways will be integrated throughout the study.

In Phase I, a cross-sectional survey will be conducted among adolescents aged 11–15 years attending public and private schools in Chandigarh and Mohali. The survey will assess the prevalence and patterns of ACEs, mental health outcomes, psychosocial wellbeing, perceived stress, sleep quality, screen-time behaviours, and behavioural and biological risk factors for non-communicable diseases (NCDs).

Phase II will comprise a qualitative exploration using focus group discussions (FGDs) and key informant interviews (KIIs) with adolescents, parents, teachers, school counsellors, child protection professionals, and mental health experts. This phase will seek to understand the lived experiences of adolescents exposed to adversity, identify contextual determinants of mental health and resilience, and explore barriers and facilitators to the implementation of trauma-informed school interventions.

Findings from the quantitative and qualitative phases will be integrated to inform Phase III, which involves the development of a culturally adapted Trauma-Informed Care and Social-Emotional Learning (TIC–SEL) intervention. The intervention will be developed using the ADDIE (Analysis, Design, Development, Implementation, and Evaluation) framework and will incorporate evidence-based strategies for emotional regulation, resilience building, stress management, coping skills enhancement, peer support, and trauma-sensitive educational practices. [10]

In Phase IV, a quasi-experimental pre–post intervention study will be conducted among adolescents reporting exposure to one or more ACEs. Eligible participants will receive the school-based TIC–SEL intervention delivered by trained psychologists in collaboration with teachers and school counsellors. The effectiveness of the intervention will be evaluated by comparing baseline and post-intervention outcomes, including emotional and behavioural wellbeing, depressive symptoms, perceived stress, sleep quality, school functioning, and behavioural risk factors for NCDs.

Phase IV is a single-arm, non-randomised evaluation: all eligible adolescents reporting one or more ACEs are offered the intervention, there is no concurrent control group and no allocation sequence, allocation concealment or allocation ratio applies. The study is open-label — participants, intervention deliverers, teachers and school counsellors are necessarily aware of intervention receipt, and outcome assessment is not blinded. To limit measurement bias, outcome instruments will be self-completed by participants under standardised conditions rather than administered by intervention facilitators, and data entry and analysis will be undertaken by personnel not involved in intervention delivery. The rationale for not randomising is set out in the Discussion.

The sequential mixed-methods approach will enable comprehensive assessment of the burden and consequences of ACEs, facilitate the development of a contextually relevant intervention, and generate evidence regarding the feasibility and effectiveness of trauma-informed school-based strategies for improving adolescent mental health and reducing future health risks.

### Study Setting

The study will be conducted in public and private schools located in cities of Chandigarh and Mohali (Punjab) North India. Both urban and rural/peri-urban schools will be included to ensure representation of diverse socio-demographic contexts.

### Participant Eligibility

#### Inclusion and Exclusion Criteria

School-going adolescents enrolled in selected public and private schools of Chandigarh and Mohali will be eligible for participation in the study. Eligible participants must be aged 11–15 years, able to understand and respond to questionnaires in Hindi, Punjabi, or English, and provide written assent along with parental/guardian consent.

For the intervention phase, adolescents reporting exposure to one or more Adverse Childhood Experiences (ACEs) during the baseline assessment and willing to participate in the intervention sessions will be eligible for enrolment.

Adolescents will be excluded from participation if they or their parents/guardians are unable or unwilling to provide informed assent or consent. Students with severe cognitive or developmental impairments that preclude meaningful participation in study assessments will also be excluded. In addition, adolescents identified as experiencing an acute psychiatric emergency, including active suicidal ideation, severe self-harm risk, psychosis, or other conditions requiring immediate specialist intervention, will not be enrolled in the intervention component and will instead be referred through the study safeguarding and mental health referral pathway for appropriate clinical care. Adolescents who are absent during both the baseline assessment and subsequent make-up visits will also be excluded from analysis.

For ethical reasons, students identified as experiencing ongoing abuse, severe psychological distress, or significant mental health concerns during the study will continue to receive counselling support and referral services irrespective of their participation in the intervention evaluation.

#### Sample Size Calculation

The sample size was calculated to estimate the prevalence of Adverse Childhood Experiences (ACEs) among school-going adolescents in Chandigarh and Mohali. Sample size estimation for a single population proportion was performed using the formula recommended by Lwanga and Lemeshow for prevalence studies [11].

Based on a large population-based study of self-reported ACEs among adolescents, in which 22.4% reported four or more ACEs, the expected prevalence of adolescents reporting four or more ACEs was taken as 22% [12]. A confidence level of 95% and an absolute precision of 3% were considered.

Using the formula:

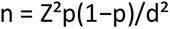

where *p* represents the expected prevalence (22%), *d* the desired precision (3%), and *Z* the standard normal deviate corresponding to a 95% confidence level (1.96), the minimum sample size under simple random sampling was estimated to be 733 adolescents. Allowing for a non-response rate of approximately 10%, this was increased to 814 adolescents.

As the study employs a multistage school-based cluster sampling design, a design effect of 2.0 was applied to account for clustering within schools, increasing the required sample size to 1,628 participants. The final sample size was rounded up to 1,632 to allow equal allocation of 102 adolescents to each of the 16 participating schools.

Accordingly, approximately 1,632 adolescents will be recruited from 16 public and private schools across Chandigarh and Mohali using multistage stratified random sampling. This sample size will provide sufficient precision for estimating ACE prevalence and adequate statistical power to examine associations between ACE exposure, mental health outcomes, and behavioural and biological risk factors for non-communicable diseases.

The sample size for the intervention phase was estimated based on the expected prevalence of ACE exposure. Assuming that approximately 30% of adolescents will report one or more ACEs, nearly 490 adolescents are expected to be eligible for intervention. All of these approximately 490 adolescents will receive the Trauma-Informed Care and Social-Emotional Learning (TIC–SEL) intervention and will undergo pre- and post-intervention assessment.

### Sampling Strategy

The adequacy of this sample for the evaluation objective was verified separately. For a paired comparison of SDQ total difficulties scores between baseline and three months post-intervention, assuming a standard deviation of 6.0 points, a correlation of 0.5 between paired measurements, two-sided alpha of 0.05 and 90% power, 100 participants are required to detect a change of 2.0 points (Cohen’s d ≈ 0.29). Allowing for a design effect of 2.0 for clustering within schools and up to 25% loss to follow-up, a minimum of 267 adolescents is required (100 × 2.0 ÷ 0.75); the planned intervention sample of approximately 490 adolescents therefore provides adequate power to detect a small-to-moderate improvement in the primary outcome, and sufficient precision for the secondary outcomes.

A multistage stratified random sampling strategy will be employed to ensure adequate representation of adolescents from diverse geographical and educational settings in Chandigarh and Mohali. The sampling framework has been designed to capture variations in exposure to adverse childhood experiences (ACEs) across urban and rural/peri-urban populations, as well as across public and private school systems.

In the first stage, the study area will be stratified according to geographical location (urban versus rural/peri-urban) and city (Chandigarh versus Mohali). This stratification will ensure balanced representation from both administrative regions and settlement types. Approximately half of the study sample (n ≈ 816) will be recruited from urban areas and the remaining half (n ≈ 816) from rural/peri-urban areas.

In the second stage, each geographical stratum will be further stratified by type of school (public versus private). Within each city-specific stratum (e.g., urban Chandigarh, urban Mohali, rural/peri-urban Chandigarh, rural/peri-urban Mohali), four schools will be selected, comprising two public and two private schools. Schools will be selected using simple random sampling from a sampling frame obtained from the respective education departments. Consequently, a total of 16 schools (8 public and 8 private) will be included in the study.

In the third stage, students will be recruited from the selected schools using stratified random sampling. Approximately 102 adolescents will be enrolled from each school, yielding a total sample size of approximately 1,632 participants. Within each school, recruitment will be stratified by gender to ensure balanced representation of boys and girls across the adolescent age spectrum.

This sampling strategy is expected to provide a representative sample of school-going adolescents from Chandigarh and Mohali while ensuring sufficient statistical power to estimate the prevalence of ACEs and examine their associations with mental health outcomes and behavioural and biological risk factors for non-communicable diseases.

### Timeframe and Implementation Strategy

The study will be conducted over a period of 36 months (3 years) from January 2026 to December 2028 and follows a sequential mixed-methods design comprising four phases, preceded by preparatory activities and followed by data analysis and dissemination: Phase I, quantitative baseline assessment; Phase II, qualitative exploration; Phase III, intervention development; and Phase IV, intervention implementation and evaluation (staff training, intervention delivery and post-intervention reassessment). The protocol was prospectively registered on 6 October 2025, following ethics approval on 27 September 2025. Preparatory activities were completed between January and March 2026; recruitment of adolescents from participating schools commenced on 16^th^ April 2026 and is expected to be completed by 30^th^ June 2027; baseline data collection is currently under way. Intervention development (Phase III) and staff training (the first component of Phase IV) will be completed after baseline assessment and before the start of intervention delivery in each school. Intervention delivery is scheduled for July 2027 to March 2028, with post-intervention reassessment three months after completion of the intervention in each school, and analysis and dissemination through December 2028. At the time of submission, the study has not generated any results; all data collection is expected to be completed by June 2028, and results are expected to be available from December 2028. The schedule of enrolment, interventions and assessments for individual participants is presented in Fig 1, following the SPIRIT recommendations. [13]

### Preparatory Activities

The preparatory phase, completed between January and March 2026, focused on obtaining all necessary regulatory approvals and establishing the study infrastructure. Approval was obtained from the Institutional Ethics Committee (IEC) on 27 September 2025, the study was registered with the Clinical Trials Registry–India (CTRI) on 6 October 2025, and permissions were obtained from the District Education Offices (DEOs) of Chandigarh and Mohali. A multistage stratified sampling framework was finalised, and participating schools were selected to ensure representation across urban and rural/peri-urban areas and public and private schools. Project personnel, including research scientists, technical support staff, and data management personnel, were recruited and trained. Study instruments, standard operating procedures, data collection platforms, and safeguarding protocols were finalised during this phase.

### Phase I: Quantitative Assessment

A cross-sectional baseline survey is being conducted among approximately 1,632 adolescents aged 11–15 years enrolled in selected schools. Following parental consent and student assent, participants will undergo comprehensive assessment using validated instruments. Data collection will include measurement of adverse childhood experiences (ACE questionnaire), emotional and behavioural difficulties (SDQ), depressive symptoms (PHQ-9), perceived stress (PSS-10), sleep quality (Child and Adolescent Sleep Checklist), screen-time behaviours (QueST), and behavioural and biological risk factors for non-communicable diseases using the WHO STEPS framework.

### Phases II and III: Qualitative Exploration and Intervention Development

Qualitative data collection will be undertaken through in-depth interviews and focus group discussions involving adolescents with ACE exposure, parents, teachers, school counsellors, child protection professionals, mental health experts, and healthcare providers. Findings from the quantitative and qualitative phases will be integrated to identify priority needs, contextual determinants, barriers, and facilitators. Using the ADDIE framework (Analysis, Design, Development, Implementation, and Evaluation), a culturally adapted Trauma-Informed Care and Social-Emotional Learning (TIC–SEL) intervention package will be developed and validated.

### Phase IV (a): Staff Training

Before intervention delivery begins in each school, capacity-building workshops will be conducted for teachers, school counsellors, psychologists, and project staff. Training will focus on trauma-informed care principles, social-emotional learning strategies, identification of ACE-related distress, safeguarding procedures, referral pathways, and standardized delivery of intervention components. Intervention manuals and implementation guides will be finalized during this phase. Participating teachers and school counsellors will rate their capacity to manage ACE-related issues on a 0–10 Likert scale before training, immediately after training, and at the 3-month post-intervention assessment.

### Phase IV (b): Intervention Delivery

Adolescents identified as having experienced one or more ACEs will participate in the TIC–SEL intervention. The intervention will be delivered over 8–12 weeks in participating schools by trained psychologists in collaboration with teachers and school counsellors. Intervention components will include emotional regulation training, stress management, coping skills development, peer-support activities, trauma-sensitive educational practices, individual counselling for high-risk adolescents (at least six sessions per adolescent), and parental sensitization on supporting adolescents with ACEs. School-wide awareness activities will also be conducted to promote a supportive and trauma-sensitive environment.

### Phase IV (c): Post-intervention Reassessment

Three months following completion of the intervention, participants will undergo repeat assessment using the same tools administered at baseline. The primary outcome will be change in emotional and behavioural difficulties (SDQ). Secondary outcomes will include depressive symptoms (PHQ-9), perceived stress, sleep quality, school functioning, screen-time behaviours, and behavioural risk factors for non-communicable diseases. Process evaluation measures including acceptability, fidelity, feasibility, and participant satisfaction will also be assessed.

## Data Analysis and Dissemination

Quantitative and qualitative data analysis will be conducted concurrently. Quantitative analyses will include prevalence estimation, regression modelling, and pre-post intervention comparisons. Qualitative data will undergo thematic analysis to explore implementation experiences and contextual influences. Findings will be disseminated through peer-reviewed publications, policy briefs, stakeholder workshops, and presentations at national and international conferences.

## Data Collection Procedures

Following approval from the Institutional Ethics Committee, District Education Offices, and participating schools, data collection will be undertaken by trained research staff using standardized procedures. School administrators, teachers, parents, and students will be sensitized regarding the study objectives, procedures, and ethical safeguards.

Information sheets and informed consent forms will be distributed to parents or legal guardians through participating schools. Written parental consent and adolescent assent will be obtained prior to enrolment. Participation will be voluntary, and confidentiality of responses will be emphasized throughout the study.

Baseline assessments will be conducted during school hours in a private and supportive environment to facilitate honest reporting of sensitive experiences. Participants will complete a structured questionnaire comprising socio-demographic information, adverse childhood experiences, mental health measures, perceived stress, sleep quality, screen-time behaviours, and behavioural risk factors for non-communicable diseases. Trained research personnel will be available to clarify questions and provide support where required.

Anthropometric measurements, including height and weight, will be obtained using calibrated equipment following standard protocols. Body mass index (BMI) will be calculated using age- and sex-specific criteria. Blood pressure measurements will be recorded using validated digital devices after an appropriate period of rest, with measurements performed by trained personnel according to standardized procedures.

Upon completion of baseline assessments, adolescents reporting one or more adverse childhood experiences will be screened for eligibility for the intervention phase. Eligible participants will be invited to participate in the school-based Trauma-Informed Care and Social-Emotional Learning (TIC– SEL) intervention. Appropriate safeguarding procedures, counselling support, and referral mechanisms will be activated for participants identified as experiencing significant psychological distress, ongoing abuse, or other concerns requiring professional attention.

Follow-up assessments will be conducted three months after completion of the intervention using the same instruments administered at baseline. To promote complete follow-up, assessment dates will be fixed in advance with each school and communicated to parents; adolescents absent on the scheduled day will be assessed during a make-up window of up to two weeks, with up to three attempts made through the school counsellor. Participants who change school or leave during the study period will be contacted through the guardian contact details recorded at enrolment and offered assessment at a mutually convenient time. Reasons for non-completion will be recorded for every participant not assessed at follow-up. These assessments will evaluate changes in emotional and behavioural wellbeing, depressive symptoms, perceived stress, sleep quality, school functioning, and behavioural risk factors for non-communicable diseases. To ensure data quality, all research staff will undergo training prior to data collection, and regular monitoring and verification procedures will be implemented throughout the study.

## Instruments and Data Collection Methods

Data will be collected using a combination of validated quantitative instruments, anthropometric measurements, and qualitative methods. All instruments selected for the study have been widely used in adolescent populations and have demonstrated acceptable psychometric properties.

Adverse childhood experiences will be assessed using the Pediatric ACEs and Related Life-events Screener (PEARLS), which captures exposure to abuse, neglect, household dysfunction, and other childhood adversities. [14] Mental health outcomes will be assessed using the Strengths and Difficulties Questionnaire (SDQ), the Patient Health Questionnaire-9 (PHQ-9), and the Perceived Stress Scale (PSS-10). [15–18] Sleep quality and sleep-related problems will be evaluated using the Child and Adolescent Sleep Checklist (CASC) [19], while screen-time behaviours will be assessed using the Questionnaire for Screen Time of Adolescents (QueST). [20] School functioning will be assessed using school attendance records, classroom behaviour reports, and teacher ratings of students’ concentration and social adjustment. The capacity of teachers and school counsellors to manage ACE-related issues will be assessed using a simple self-rated 11-point Likert scale (0 = no capacity, 10 = full capacity). Staff will rate their capacity to recognise ACE-related distress, respond in a trauma-informed manner, support affected students, and refer students through the safeguarding pathway. All instruments have established reliability and validity in adolescent populations, and the SDQ, PHQ-9 and PSS-10 have been used previously in Indian adolescent samples. Where a validated Hindi or Punjabi version exists it will be used; where none exists, the instrument will be translated and back-translated by bilingual experts and pre-tested with 30 adolescents in a non-participating school, with internal consistency (Cronbach’s alpha) reported for each translated instrument at baseline. Outcomes, instruments and assessment time points are summarised in Table 1.

**Table 1.** Primary and Secondary Outcomes, Instruments, Data Collection Methods and Time of Assessment.

| Outcome Level | Outcome Measure | Instrument / Data Collection Method | Time of Assessment |
| --- | --- | --- | --- |
| **Primary Outcome** | Emotional & Behavioural well being | Strength And Difficulties Questionnaire | Baseline; 3 months post-intervention |
| **Secondary Outcomes** | Depressive symptoms (key secondary outcome) | Patient Health Questionnaire-9 (PHQ-9) | Baseline; 3 months post-intervention |
|  | Perceived stress | Perceived Stress Scale-10 (PSS-10) | Baseline; 3 months post-intervention |
|  | Adverse childhood experiences | PEARLS Questionnaire | Baseline |
|  | Sleep quality and sleep problems | Child and Adolescent Sleep Checklist (CASC) | Baseline; 3 months post-intervention |
|  | Screen-time behaviours | Questionnaire for Screen Time of Adolescents (QueST) | Baseline; 3 months post-intervention |
|  | Tobacco use | Modified WHO STEPS questionnaire | Baseline; 3 months post-intervention |
|  | Alcohol use | Modified WHO STEPS questionnaire | Baseline; 3 months post-intervention |
|  | Dietary habits | Modified WHO STEPS questionnaire | Baseline; 3 months post-intervention |
|  | Physical activity | Modified WHO STEPS questionnaire | Baseline; 3 months post-intervention |
|  | Body mass index (BMI) | Height and weight measurements by trained personnel | Baseline; 3 months post-intervention |
|  | Blood pressure | Standardized BP measurement using validated devices | Baseline; 3 months post-intervention |
|  | School functioning | School attendance records, classroom behaviour reports, and teacher ratings of concentration and social adjustment | Baseline; 3 months post-intervention |
|  | Capacity of school staff to manage ACE-related issues | Self-rated 0–10 Likert scale completed by teachers and school counsellors | Before training; immediately after training; 3 months post-intervention |
| **Sociodemographic Variables** | Age, gender, family structure, socioeconomic status, school characteristics | Structured questionnaire | Baseline |
| **Process Evaluation** | Acceptability, feasibility, fidelity, participant satisfaction | Attendance records, structured feedback forms, observation checklists, interviews | During and post-intervention |
| **Qualitative Component** | Experiences of adversity, mental health concerns, school support systems, intervention acceptability, barriers and facilitators | Focus Group Discussions and Key Informant Interviews | Intervention development and process evaluation phases |

Abbreviations: ACEs, Adverse Childhood Experiences; PEARLS, Pediatric ACEs and Related Life-events Screener; SDQ, Strengths and Difficulties Questionnaire; PHQ-9, Patient Health Questionnaire-9; PSS-10, Perceived Stress Scale-10; CASC, Child and Adolescent Sleep Checklist; QueST, Questionnaire for Screen Time of Adolescents; WHO STEPS, World Health Organization STEPwise approach to NCD risk factor surveillance.

Behavioural and biological risk factors for non-communicable diseases (NCDs) will be assessed using a modified WHO STEPwise approach to NCD risk factor surveillance. [21] This will include information on tobacco and alcohol use, dietary habits, physical activity, anthropometric measurements (height, weight, and body mass index), and blood pressure. Sociodemographic information including age, gender, family structure, socioeconomic status, and school characteristics will also be collected.

Qualitative data will be obtained through focus group discussions (FGDs) and key informant interviews (KIIs). FGDs will be conducted with adolescents, parents, teachers, and school counsellors, while KIIs will be undertaken with psychologists, psychiatrists, Child Welfare Committee members, child protection professionals, and education administrators. Discussions will explore experiences of adversity, mental health concerns, existing school support systems, acceptability of trauma-informed interventions, and implementation barriers and facilitators. All interviews and discussions will be audio-recorded, transcribed verbatim, and analysed using thematic analysis. [22]

## Qualitative Component

The qualitative component will be undertaken during Phase II of the study to gain an in-depth understanding of the lived experiences of adolescents exposed to adverse childhood experiences (ACEs), explore contextual factors influencing mental health and resilience, and inform the development of a culturally appropriate Trauma-Informed Care and Social-Emotional Learning (TIC– SEL) intervention.

A combination of Focus Group Discussions (FGDs) and Key Informant Interviews (KIIs) will be conducted using semi-structured interview guides developed from the literature review, findings from the quantitative phase, and expert consultation. FGDs will be conducted separately with adolescents, parents, teachers, and school counsellors to explore perceptions of childhood adversity, mental health concerns, coping mechanisms, available support systems, and perspectives regarding trauma-informed interventions within schools.

Key Informant Interviews will be conducted with psychologists, psychiatrists, Child Welfare Committee members, child protection professionals, and education administrators. These interviews will provide insights into existing child mental health and protection services, referral pathways, implementation challenges, policy considerations, and opportunities for integrating trauma-informed approaches within school health programmes.

The qualitative enquiry will focus on understanding experiences of adversity, mental health concerns among adolescents, existing school and community support systems, perceived needs for psychosocial support, acceptability of trauma-informed care interventions, and potential barriers and facilitators to implementation. Particular attention will be given to identifying culturally relevant intervention components and strategies to enhance feasibility, sustainability, and scalability within the Indian school context.

All interviews and discussions will be conducted by trained qualitative researchers in a private setting and in the preferred language of participants (Hindi, Punjabi, or English). Interviews will be audio-recorded with participant consent, transcribed verbatim, and translated into English where necessary. Data collection will continue until thematic saturation is achieved.

Qualitative data will be analysed using Braun and Clarke’s six-step thematic analysis approach. Two researchers will independently review transcripts, generate initial codes, identify emerging themes, and develop a coding framework. Themes will be refined through iterative discussions among the research team to ensure credibility and trustworthiness. Findings from the qualitative phase will be triangulated with quantitative results and will directly inform the development, adaptation, and implementation of the TIC–SEL intervention.

## Intervention Development

The Trauma-Informed Care and Social-Emotional Learning (TIC–SEL) intervention will be developed using the ADDIE [10] instructional design framework, comprising the phases of Analysis, Design, Development, Implementation, and Evaluation. This systematic approach will ensure that the intervention is evidence-based, contextually relevant, and responsive to the needs of adolescents exposed to adverse childhood experiences (ACEs).

During the Analysis phase, findings from the literature review, quantitative assessment, and qualitative exploration will be synthesized to identify priority mental health concerns, psychosocial needs, contextual determinants, and existing gaps in school-based support systems. These findings will guide the conceptualization of intervention components and delivery strategies.

In the Design phase, learning objectives, intervention content, session structure, pedagogical approaches, and implementation strategies will be developed. Particular emphasis will be placed on integrating principles of trauma-informed care with social-emotional learning to address both the psychological consequences of adversity and the development of resilience-promoting skills.

The Development phase will involve preparation of intervention materials, including facilitator manuals, teacher guides, student workbooks, counselling resources, activity sheets, and audiovisual learning materials. Draft materials will undergo expert review and iterative refinement to ensure content validity, cultural appropriateness, and feasibility for implementation within school settings.

During the Implementation phase, the intervention package will be pilot-tested in selected schools. Feedback from students, teachers, counsellors, and implementation staff will be used to refine the intervention prior to wider rollout.

Finally, the Evaluation phase will include expert validation, field testing, and assessment of feasibility, acceptability, and implementation readiness. Findings from this phase will inform final modifications to the intervention package.

The intervention curriculum will address key domains identified through the needs assessment, including emotional awareness, emotional regulation, resilience building, stress management, problem-solving skills, communication and interpersonal skills, positive relationships, help-seeking behaviours, and trauma-informed coping strategies.

## Intervention Implementation

The TIC–SEL intervention will be delivered over a period of 8–12 weeks by trained psychologists in collaboration with school counsellors and teachers. Intervention sessions will include individual counselling sessions for high-risk adolescents (at least six sessions per adolescent), school-wide awareness activities to create a trauma-sensitive environment, and parental sensitization on supporting adolescents with ACEs. Session titles, learning objectives, activities and materials will be specified in the facilitator manual, and the intervention will be reported in full using the Template for Intervention Description and Replication (TIDieR) checklist. [23]

Sessions will be delivered by clinical psychologists holding an MPhil or PhD in Clinical Psychology with prior experience of working with children and adolescents, supported by school counsellors and trained teachers. All deliverers will complete the structured training programme described under Phase IV (a) before delivering any session, and will receive fortnightly supervision from the study psychiatrist and clinical psychologist. Fidelity will be assessed using a session-specific checklist completed by the facilitator and verified by a supervisor on a random sample of sessions.

To support adherence, sessions will be scheduled within school hours in consultation with the school administration, attendance will be recorded at each session, and participants who miss a session will be offered a make-up session before the next module begins. Participants absent from three or more consecutive sessions will be contacted through the school counsellor to identify and address barriers to participation.

Intervention content may be adapted in delivery style, language or pacing to suit the comprehension level of a group, provided the core components are retained; any such adaptation will be documented. A participant will be discontinued from intervention sessions if they develop an acute psychiatric emergency, express a wish to withdraw, or if the study psychiatrist judges that continued participation is not in the participant’s interest. Discontinuation from the intervention does not entail withdrawal from follow-up assessment unless the participant or guardian withdraws consent, and clinical care and referral continue in all cases. The intervention will adopt a multi-level approach targeting students, teachers, families, and the broader school environment to create a trauma-sensitive ecosystem that promotes resilience and emotional wellbeing.

Core intervention components will include social-emotional learning content focusing on emotional regulation, resilience, stress management, communication skills, and adaptive coping strategies. Adolescents identified as high risk during screening or intervention delivery will receive individual counselling (at least six sessions per adolescent) and referral support as appropriate.

To strengthen institutional capacity, teacher sensitization and capacity-building workshops will be conducted before intervention delivery, as part of the Phase IV staff training, to enhance understanding of ACEs, trauma-informed practices, and strategies for supporting vulnerable students within classroom settings. Parental sensitization sessions will be organized to improve awareness of adolescent mental health, strengthen supportive caregiving practices, and promote positive family communication.

Additional components will include peer-support activities designed to foster social connectedness and help-seeking behaviours, as well as school-wide awareness initiatives aimed at promoting a safe, inclusive, and trauma-sensitive school environment. Appropriate safeguarding procedures and referral pathways will be integrated throughout the intervention.

## Outcome Measures

The primary outcome of the study will be the change in emotional and behavioural difficulties measured using the Strengths and Difficulties Questionnaire (SDQ) total difficulties score, from baseline to three months post-intervention. Consistent with Objective 3 of the funded protocol, the SDQ total difficulties score is the single primary outcome. Depressive symptoms (PHQ-9), which were listed alongside the SDQ in the protocol’s implementation chart, are designated the key secondary outcome and will be reported first among secondary outcomes.

Secondary outcomes will include changes in depressive symptoms (PHQ-9), perceived stress, sleep quality, screen-time behaviours, school functioning (school attendance records, classroom behaviour reports, and teacher-rated concentration and social adjustment), capacity of teachers and school counsellors to manage ACE-related issues (self-rated 0–10 Likert scale), and behavioural risk factors for non-communicable diseases, including unhealthy dietary practices, physical inactivity, tobacco use, and alcohol use.

In addition to effectiveness outcomes, a process evaluation will be conducted to assess implementation fidelity, intervention reach and participation, acceptability among stakeholders, feasibility of delivery within school settings, and participant satisfaction. These process indicators will provide critical information regarding the scalability and sustainability of the intervention in real-world educational settings.

## Process Evaluation

A comprehensive process evaluation will be conducted alongside the intervention to assess the quality, feasibility, acceptability, and fidelity of implementation of the Trauma-Informed Care and Social-Emotional Learning (TIC–SEL) intervention. The process evaluation will provide critical information regarding how the intervention is delivered, received, and integrated within participating schools, thereby facilitating interpretation of intervention outcomes and informing future scale-up efforts.

Key process indicators will include intervention fidelity, dose delivered, dose received, participant engagement, adherence of teachers and counsellors to intervention protocols, and overall acceptability among students, parents, teachers, counsellors, and other stakeholders. Intervention fidelity will assess the extent to which intervention components are delivered as intended, while dose delivered and dose received will evaluate the quantity of intervention activities provided and the level of participant exposure to those activities. Participant engagement will be assessed through attendance, active participation in sessions, completion of assigned activities, and involvement in peer-support initiatives.

Teacher and counsellor adherence will be evaluated by examining compliance with intervention guidelines, delivery of planned activities, and integration of trauma-informed practices within the school environment. Stakeholder acceptability will be explored through perceptions of relevance, usefulness, appropriateness, and satisfaction with the intervention.

Multiple data sources will be used for process evaluation, including structured observation checklists, facilitator logs, attendance records, intervention monitoring forms, participant satisfaction surveys, and qualitative interviews with students, teachers, school counsellors, parents, and intervention facilitators. Qualitative data will further explore perceived benefits, implementation challenges, contextual influences, and recommendations for improvement.

Findings from the process evaluation will be triangulated with quantitative outcome data to identify factors influencing intervention effectiveness and to assess the feasibility and scalability of implementing trauma-informed school-based mental health interventions within the Indian educational context.

## Safeguarding and Referral Protocol

Given the sensitive nature of assessing Adverse Childhood Experiences (ACEs), a comprehensive safeguarding and referral protocol will be implemented throughout all phases of the study to ensure participant safety, psychological wellbeing, and compliance with applicable Indian child protection laws and policies. All study personnel will receive training in child safeguarding, trauma-informed approaches, confidentiality, mandatory reporting requirements, and referral procedures prior to initiation of data collection.

During assessments and intervention activities, participants will be screened for indicators of immediate risk, including suicidal ideation, self-harm behaviours, severe depressive symptoms, acute psychological distress, ongoing physical, emotional, or sexual abuse, neglect, exploitation, trafficking, child marriage, substance misuse, and other circumstances that may place the child at risk of harm. Any participant identified as being at immediate risk will be promptly evaluated and referred according to the safeguarding pathway.

A tiered referral system will be utilized. At the first level, students experiencing mild to moderate emotional or psychosocial concerns will receive support through school counsellors or designated mental health personnel or school Principal within the school system. At the second level, participants requiring specialized assessment or counselling will be referred to the project psychologist or trained mental health professionals associated with the study. At the third level, adolescents presenting with severe mental health concerns, suicidal risk, significant self-harm behaviours, severe depression, psychosis, or other psychiatric emergencies will be referred to Psychiatry or Clinical Psychology services at participating tertiary care institutions for comprehensive evaluation and management.

All cases involving suspected abuse, neglect, abandonment, exploitation, trafficking, child labour, children living without parental care, children affected by violence, or other situations where a child may be considered a Child in Need of Care and Protection (CNCP) will be managed in accordance with the provisions of the Juvenile Justice (Care and Protection of Children) Act, 2015 [24], and the operational guidelines of Mission Vatsalya [25]. Such cases will be referred to the appropriate authorities, including the Child Welfare Committee (CWC), District Child Protection Unit (DCPU), Childline services (1098), Special Juvenile Police Unit (SJPU), or other designated child protection agencies, as appropriate.

Cases involving disclosure of sexual abuse, sexual assault, sexual harassment, or online sexual exploitation will be managed in accordance with the Protection of Children from Sexual Offences (POCSO) Act, 2012 [26]. Mandatory reporting obligations under the POCSO Act will be followed, and appropriate referrals will be made to child protection and legal authorities while ensuring the child’s safety and psychological support.

Where adolescents are identified as being involved in behaviours that may bring them into contact with the justice system, support and referrals will be provided in accordance with the provisions of the Juvenile Justice Act relating to Children in Conflict with Law (CCL) [24]. Appropriate coordination may be undertaken with the Juvenile Justice Board (JJB), child protection services, and mental health professionals to ensure a child-centred and rehabilitative approach.

A crisis management protocol will be available during all assessments, interviews, focus group discussions, and intervention sessions. Participants requiring urgent mental health care or protection services will receive immediate attention, including parental or guardian notification where appropriate, emergency referral, and follow-up support. Confidentiality will be maintained to the greatest extent possible; however, participants will be informed that confidentiality may be breached when disclosure is necessary to protect the child from harm or to comply with legal reporting obligations.

Adverse events will be defined, ascertained and reported separately from safeguarding actions. Anticipated study-related harms include acute distress during ACE screening or intervention sessions, re-traumatisation on discussing adverse experiences, unintended disclosure of sensitive information to peers or family members, deterioration in mood or emerging suicidal ideation during the intervention period, and stigmatisation arising from participation in a targeted intervention group. Events will be ascertained through facilitator observation and session logs, direct participant report, teacher and counsellor reporting, and review of scheduled assessments; an adverse event form will be completed for each event recording its nature, date, severity, presumed relatedness to study procedures, action taken and outcome. Serious adverse events — including suicide attempt, hospitalisation, and any event resulting in withdrawal from the intervention — will be reported to the Principal Investigator within 24 hours and to the Institutional Ethics Committee within seven days, in accordance with institutional requirements. Aggregate adverse event data will be reviewed at monthly project meetings and reported in full in the results publication irrespective of statistical significance.

The safeguarding protocol is intended to ensure that participation in the study does not increase risk to adolescents and that children identified as vulnerable receive timely access to appropriate mental health, social welfare, and child protection services.

## Data Management

All study data will be collected using standardized paper-based questionnaires and data collection forms by trained research personnel. Each participant will be assigned a unique study identification number to ensure confidentiality and facilitate de-identification of study records. Personal identifiers and research data will be maintained separately and linked only through unique participant codes.

Completed questionnaires and data collection forms will be reviewed on the day of collection to ensure completeness and accuracy. Data will subsequently be entered into a study database by designated research personnel. Regular data verification and quality assurance procedures, including consistency checks, review of missing data, and validation of entered records against source documents, will be undertaken to ensure data accuracy and integrity.

Access to study records will be restricted to authorized members of the research team. Data quality will be monitored through monthly review of a random 10% sample of completed instruments against the electronic database by an investigator not involved in that school’s data collection, with discrepancy rates recorded and corrective action documented. Monitoring visits to each participating school will be conducted at least once per data collection cycle. These monitoring procedures are internal to the study team and independent of the funding agency. Hard-copy questionnaires, consent forms, interview transcripts, and other study documents will be stored in locked almirahs within secure institutional premises. All study personnel will be trained in maintaining participant confidentiality and ethical handling of sensitive information.

Qualitative interviews and focus group discussions will be audio-recorded with participant consent, transcribed verbatim, and de-identified prior to analysis. Any information that could directly identify participants will be removed from transcripts and reports.

All data will be handled in accordance with institutional ethical guidelines and applicable regulatory requirements. Study findings will be reported only in aggregate form, and no information that could identify individual participants, schools, or communities will be disclosed in publications, presentations, or reports.

## Statistical Analysis

### Quantitative Analysis

Data analysis will be performed using IBM SPSS Statistics version 21 (IBM Corp., Armonk, NY, USA) and R version 4.5.2 (R Foundation for Statistical Computing, Vienna, Austria) run in RStudio. Prior to analysis, data will be examined for completeness, consistency, normality, and outliers. Continuous variables will be summarized using means and standard deviations or medians and interquartile ranges, as appropriate. Categorical variables will be summarized using frequencies and percentages.

For Objective 1, the prevalence and distribution of adverse childhood experiences (ACEs) will be estimated overall and across sociodemographic subgroups. The prevalence of individual ACE categories and cumulative ACE scores will be reported with corresponding 95% confidence intervals.

For Objective 2 (Objectives 2a and 2b), bivariate analyses will be conducted to examine associations between ACE exposure and mental health outcomes, psychosocial variables, and behavioural and biological risk factors for non-communicable diseases (NCDs). Chi-square tests or Fisher’s exact tests will be used for categorical variables, while independent t-tests and one-way analysis of variance (ANOVA) will be used for comparisons involving normally distributed continuous variables. Non-parametric alternatives, including Mann–Whitney U and Kruskal–Wallis tests, will be employed when assumptions of normality are not met.

Multivariable analyses will be conducted to identify independent predictors of ACE exposure and associated health outcomes while adjusting for potential confounders. Logistic regression models will be used for binary outcomes, whereas multiple linear regression analyses will be used for continuous outcome variables. Adjusted odds ratios (AORs), regression coefficients, and 95% confidence intervals will be reported.

For Objective 3, intervention effectiveness will be assessed by comparing baseline and post-intervention outcomes among adolescents participating in the Trauma-Informed Care and Social-Emotional Learning (TIC–SEL) intervention. Changes in continuous outcomes such as SDQ scores, PHQ-9 scores, perceived stress, and sleep quality will be summarised descriptively and compared using paired t-tests for normally distributed data and Wilcoxon signed-rank tests for non-normally distributed data as unadjusted analyses. The primary analysis will use mixed-effects models accounting for within-subject correlation and clustering by school, as described below. Subgroup analyses may be conducted to explore differential intervention effects according to gender, age group, school type, socioeconomic status, and ACE burden (Objective 3f).

Because participants are recruited within schools, all analyses of intervention effect will account for clustering at the school level. Mixed-effects models with a random intercept for school will be used as the primary analytic approach for continuous outcomes, with time as a fixed effect; where model assumptions are not met, cluster-robust standard errors will be applied. For all primary and secondary outcomes, the mean change with its 95% confidence interval and a standardised effect size will be reported alongside the test statistic and p-value.

The primary analysis population will comprise all adolescents who received at least one intervention session and completed a baseline assessment, analysed on an all-enrolled basis. Mixed-effects models will be fitted using all available observations under a missing-at-random assumption; a sensitivity analysis using multiple imputation will be conducted if follow-up loss exceeds 15%, and a per-protocol analysis restricted to participants attending at least 75% of sessions will be reported as a secondary analysis. Reasons for loss to follow-up will be tabulated, and baseline characteristics of completers and non-completers will be compared.

Reporting of results will follow the TREND statement for non-randomised evaluations. [27] A participant flow diagram will report the numbers screened, found eligible, consented, enrolled in the intervention, receiving each session block, assessed at follow-up and included in each analysis, with reasons for exclusion at every stage. Baseline sociodemographic characteristics, ACE exposure and outcome scores will be presented in a table for the enrolled cohort, and the denominator will be stated for every reported estimate. Adverse events will be reported in full, and all pre-specified outcomes will be reported irrespective of the direction or significance of the findings.

Subgroup analyses are exploratory and hypothesis-generating; no adjustment for multiplicity will be made, and findings will be interpreted accordingly. Change in staff capacity scores (0–10 Likert scale) across the three time points will be summarised using medians and interquartile ranges. Changes will be compared using the Wilcoxon signed-rank test for paired comparisons and the Friedman test across all three time points.

All statistical tests will be two-tailed, and statistical significance will be considered at a p-value of <0.05.

## Qualitative Analysis

Qualitative data obtained from focus group discussions and key informant interviews will be analysed using Braun and Clarke’s six-step thematic analysis approach. Audio recordings will be transcribed verbatim, translated into English where required, and reviewed for accuracy before analysis.

Two researchers will independently read and familiarize themselves with the transcripts, generate initial codes, and identify emerging themes. Codes will be grouped into broader categories and refined iteratively through discussion and comparison across transcripts. Discrepancies in coding and theme development will be resolved through consensus, with involvement of a third researcher where necessary.

Strategies to enhance trustworthiness will include investigator triangulation, maintenance of an audit trail, reflexive memo writing, and regular review of emerging themes within the research team. Findings from the qualitative component will be triangulated with quantitative results to provide a comprehensive understanding of adolescent experiences, contextual determinants of ACEs, and factors influencing the development, implementation, and effectiveness of the TIC–SEL intervention.

## Discussion

To the best of our knowledge, this study will be among the first comprehensive mixed-methods investigations in India to examine the prevalence, patterns, and consequences of Adverse Childhood Experiences (ACEs) among school-going adolescents while simultaneously developing and evaluating a culturally adapted Trauma-Informed Care and Social-Emotional Learning (TIC–SEL) intervention. The study addresses a critical evidence gap, as recent global systematic reviews have identified only one eligible study from India, highlighting the paucity of context-specific evidence on childhood adversity and its consequences among Indian adolescents.

Several strengths characterize the proposed study. First, the intervention is grounded in a strong theoretical foundation, integrating the ACE framework, toxic stress model, trauma-informed care principles, social-emotional learning, and a clearly articulated theory of change. Second, the sequential mixed-methods design combines epidemiological assessment with qualitative inquiry, enabling a comprehensive understanding of both the burden of ACEs and the contextual factors influencing implementation. Third, the intervention will be developed using the ADDIE framework and informed by adolescents, parents, teachers, school counsellors, mental health professionals, and child protection stakeholders, thereby enhancing its cultural relevance, acceptability, and feasibility.

Fourth, the study adopts a socioecological approach by targeting multiple stakeholders within the school ecosystem, including students, teachers, counsellors, and families, rather than focusing solely on individual-level behaviour change. Fifth, the study evaluates a broad range of outcomes encompassing emotional and behavioural wellbeing, depressive symptoms, perceived stress, sleep quality, school functioning, and behavioural risk factors for non-communicable diseases (NCDs). Finally, the incorporation of process evaluation will facilitate understanding of implementation fidelity, acceptability, and scalability within real-world school settings.

Unlike many intervention studies, a randomized controlled trial design was not considered ethically appropriate for the intervention phase. Once adolescents are identified as having experienced adversity and are found to be at increased risk for mental health difficulties, withholding potentially beneficial psychosocial support from a control group would raise important ethical concerns. Trauma-informed care principles emphasize timely recognition and response to adversity. Consequently, a quasi-experimental pre–post design was selected to ensure that all eligible adolescents identified with ACE exposure have access to intervention and support services while still permitting rigorous evaluation of intervention effectiveness. This approach is consistent with ethical obligations toward vulnerable children and adolescents and aligns with child rights-based and trauma-informed frameworks.

The study has important public health implications. India is home to the world’s largest adolescent population and is experiencing a growing burden of mental health disorders and behavioural risk factors for NCDs. By identifying pathways linking childhood adversity to mental health and health-risk behaviours, the study may contribute to early prevention strategies capable of reducing long-term health, social, and economic consequences associated with untreated childhood trauma.

The proposed intervention is also highly relevant to the objectives of the National Education Policy (NEP) 2020, which emphasizes holistic development, socio-emotional learning, student wellbeing, and the creation of safe and inclusive learning environments. By strengthening emotional competencies, resilience, and supportive school climates, the TIC–SEL intervention operationalizes several key recommendations of the NEP and provides a practical framework for translating policy into action within educational settings.

Similarly, the study aligns with the National Mental Health Policy of India and the School Health Programme under Ayushman Bharat, both of which advocate preventive, promotive, and school-based approaches to mental health. The study responds directly to calls for early identification of mental health concerns and strengthening psychosocial support systems for children and adolescents. Furthermore, the integration of trauma-informed care principles represents an important advancement in school mental health programming, where the impact of childhood adversity often remains under-recognized.

A key strength of the proposed intervention is its potential for scalability. The intervention utilizes existing school infrastructure and leverages teachers, school counsellors, and educational systems as implementation partners. The development of standardized manuals, training materials, implementation guides, and referral pathways will facilitate future adaptation and expansion across diverse educational settings. If found to be effective and acceptable, the TIC–SEL model could serve as a scalable framework for integration into school health and mental health programmes across India and other low- and middle-income country settings facing similar challenges.

Several limitations should be acknowledged. First, the single-arm pre–post design, although ethically justified, does not permit attribution of observed change to the intervention: maturation, regression to the mean, secular trends in adolescent mental health, and repeated exposure to the same instruments may all contribute to pre–post differences. Findings will therefore be interpreted as change associated with intervention delivery rather than as a causal effect estimate. Second, the study is open-label, and neither participants nor those delivering the intervention or assessing outcomes can be blinded, introducing the possibility of social desirability and expectation bias in self-reported outcomes. Third, exposures and most outcomes are self-reported and, in the case of ACEs, retrospectively recalled, which may lead to under-reporting of sensitive experiences despite the confidential assessment conditions. Fourth, adolescents identified as being at acute psychiatric risk are excluded from the intervention component on ethical grounds, so effects among the most severely affected group will not be estimated. Fifth, restriction to public and private schools in Chandigarh and Mohali limits generalisability to out-of-school adolescents and to other regions of India. Finally, the three-month post-intervention assessment cannot establish whether any improvements are sustained, or whether the hypothesised reduction in long-term NCD risk is realised; longer-term follow-up will be sought through subsequent funding.

Ultimately, this study is expected to generate critical evidence on the burden and consequences of ACEs among Indian adolescents and provide one of the first evaluations of a culturally adapted trauma-informed school intervention in the country. The findings may inform policy, strengthen school mental health services, enhance child protection responses, and contribute to the development of resilient, supportive educational environments capable of mitigating the long-term effects of childhood adversity.

## Ethical Approval

Ethical approval was obtained from the Institutional Ethics Committee–Human Research (IEC-HR), Dr. B.R. Ambedkar State Institute of Medical Sciences, Mohali, Punjab, India (Approval No. AIMS/IEC-HR/2025/113; dated 27 September 2025; Project ID: AIMS2025-IEC-HR-110-EM-07). Written informed consent from parents/legal guardians and assent from adolescents will be obtained before participation. The consent and assent documents state the limits of confidentiality — specifically that disclosure of ongoing abuse, sexual offences or imminent risk of harm carries a mandatory reporting obligation under the Protection of Children from Sexual Offences Act, 2012 and the Juvenile Justice (Care and Protection of Children) Act, 2015 — so that participants and guardians understand before enrolment what cannot be kept confidential. Consent is also sought separately for the use of de-identified data in future ethically approved secondary analyses; participation is not conditional on granting this optional consent. No biological specimens (such as blood, saliva or urine) will be collected; physical assessments are limited to non-invasive anthropometric and blood pressure measurements.

Any amendment to the protocol will be documented in a dated revision log, submitted to the Institutional Ethics Committee for approval before implementation, and updated in the Clinical Trials Registry–India record. Substantive amendments affecting participant procedures, risk or outcomes will be communicated to participating schools, the District Education Offices, and — where the consent or assent terms are affected — to parents/guardians and adolescents through revised information sheets and re-consent. Amendments will be reported in any resulting publication.

## Confidentiality

All study data will be de-identified at source. Unique participant identification numbers will be used in place of personal identifiers. School-level and individual-level results will be reported only in aggregate form. No information that could identify individual participants, families, schools or communities will be disclosed in publications, reports or presentations. Access to identifiable data will be restricted to authorized study personnel only

## Availability of data and materials

The minimal de-identified dataset underlying the results, together with the data dictionary and analysis code, will be deposited in a public repository and cited in the results publication. Because participants are minors reporting adverse childhood experiences, and because the schools sampled are few enough to be identifiable, variables carrying a residual risk of re-identification (school identifier, date of birth, item-level ACE responses and free-text entries) cannot be shared publicly. Requests for access to that restricted subset should be directed to the Secretary, Institutional Ethics Committee–Human Research, Dr B.R. Ambedkar State Institute of Medical Sciences, Mohali, Punjab, India, which will consider each request against the terms of the participants’ consent and applicable child-protection law. Access decisions are made by the Committee, not by the investigators, and the same conditions apply to requests from the authors’ own institution. Qualitative transcripts will not be shared in full, as the risk of deductive disclosure cannot be eliminated for a small number of identifiable schools; anonymised excerpts will be provided on request. The intervention facilitator manual, teacher guide, student workbook and training materials will be made available for non-commercial research and educational use on request.

## Access to data during the study

The Principal Investigator and named co-investigators will have access to the final dataset. There are no contractual agreements limiting investigator access to the data. The funding agency has no access to individual-level data and no role in analysis, interpretation or the decision to publish.

## Data monitoring

Given the low-risk, non-pharmacological nature of the intervention, no independent data monitoring committee has been constituted. Oversight of participant safety, adverse events and safeguarding referrals will be exercised by the Principal Investigator with the study psychiatrist and clinical psychologist at monthly project review meetings, and reported to the Institutional Ethics Committee in the annual progress report. No interim efficacy analyses are planned and no formal stopping rules apply.

Patient and public involvement: Adolescents, parents, teachers and school counsellors are involved in the design of the intervention through the Phase II qualitative component, and stakeholder feedback during pilot testing will shape the final intervention package. Adolescent and parent representatives will be invited to review lay summaries of the findings, and results will be returned to participating schools through stakeholder workshops before wider dissemination. Participants were not involved in developing the original research question or funding application.

## Data Availability

The minimal de-identified dataset underlying the results, together with the data dictionary and analysis code, will be deposited in a public repository and cited in the results publication. Because participants are minors reporting adverse childhood experiences, and because the schools sampled are few enough to be identifiable, variables carrying a residual risk of re-identification (school identifier, date of birth, item-level ACE responses and free-text entries) cannot be shared publicly. Requests for access to that restricted subset should be directed to the Secretary, Institutional Ethics Committee–Human Research, Dr B.R. Ambedkar State Institute of Medical Sciences, Mohali, Punjab, India, which will consider each request against the terms of the participants' consent and applicable child-protection law. Access decisions are made by the Committee, not by the investigators, and the same conditions apply to requests from the authors' own institution. . Qualitative transcripts will not be shared in full, as the risk of deductive disclosure cannot be eliminated for a small number of identifiable schools anonymised excerpts will be provided on request. The intervention facilitator manual, teacher guide, student workbook and training materials will be made available for non-commercial research and educational use on request.

## Author contributions

BB conceived the study, secured funding and leads the protocol development as Principal Investigator. NM and AV contributed to the design of the psychiatric and community-health components respectively. PM, DL and CP contributed to the design of the psychological assessment battery and the intervention curriculum. SB is involved in methodology and statistical planning. All authors contributed to drafting the protocol, reviewed the manuscript critically for intellectual content, and approved the final version.

## Sponsor and contact

The study sponsor is Dr B.R. Ambedkar State Institute of Medical Sciences, Mohali, Punjab. Enquiries regarding the protocol should be addressed to the corresponding author, whose contact details appear on the title page. The sponsor has no role in study design, data collection, analysis, interpretation or the decision to submit for publication.

## Authorship

Authorship of publications arising from this study will follow the International Committee of Medical Journal Editors (ICMJE) criteria. No professional writers will be used.

## Supporting Information

S1 Checklist. Completed SPIRIT 2025 checklist. Each SPIRIT 2025 item is mapped to the page of this protocol on which it is reported; items relating to randomisation, allocation concealment and blinding are marked not applicable, as Phase IV is a single-arm, open-label, non-randomised evaluation.

S2 Checklist. Reporting-guideline mapping of this protocol against the TREND statement. Maps each TREND item to the section of this protocol where it is addressed, together with the revisions made in response.

S1 File. Parent/guardian information sheet and informed consent form (English, Hindi and Punjabi).

S2 File. Adolescent assent form (English, Hindi and Punjabi).

